# DeepQCT: Predicting fragility fracture from high-resolution peripheral quantitative CT using deep learning

**DOI:** 10.1101/2024.04.01.24305147

**Authors:** Fangyuan Chen, Lijia Cui, Qiao Jin, Yushuo Wu, Jiaqi Li, Yan Jiang, Yue Chi, Ruizhi Jiajue, Wei Liu, Wei Yu, Qianqian Pang, Ou Wang, Mei Li, Xiaoping Xing, Xuegong Zhang, Weibo Xia

## Abstract

**Background:** Osteoporosis is prevalent in elderly women, which causes fragility fracture and hence increased mortality and morbidity. Predicting osteoporotic fracture risk is both clinically-beneficial and cost-effective. However, traditional tools using clinical factors and bone mineral density (BMD) fail to reflect bone microstructure. Here we aim to use high-resolution peripheral quantitative CT (HR-pQCT) images to construct deep-learning models which predict fragility fracture history in elderly Chinese women.

**Methods:** We used ChiVOS, a community-based national cohort of 2,664 Chinese elderly women. Demographic data, BMD, and HR-pQCT from 216 patients were used to construct three groups of models: BMD, pQCT-index, and DeepQCT. For DeepQCT, we used ResNet34 as classifier, and logistic regression for late fusion. Models were developed using 6-fold cross-validation in development set (90%, N=195), and tested in internal test set (10%, N=21). We applied unsupervised clustering on HR-pQCT indices to derive patient subgroups.

**Findings:** DeepQCT (best model AUC 0.86-0.94) was superior or similar to pQCT-index (best model AUC 0.8-0.93), which both outperformed BMD (best model AUC 0.54-0.78). Surprisingly, DeepQCT built from non-weight-bearing bones performed similarly to weight-bearing bones. Furthermore, two distinct patient groups were classified using HR-pQCT indices. The one with higher DeepQCT risk score showed lower volumetric BMD, bone more microarchitectural abnormalities, and had higher probability of osteoporosis and fragility fracture history.

**Interpretation:** DeepQCT scores and HR-pQCT-index permit early recognition of patients with high risk of fragility fracture. This established framework can be easily adapted for other diagnostic tasks using HR-pQCT scans, which promotes bone health management via digital medicine.

**Funding:** This research was supported by the National Natural Science Foundation of China (LC, 82100946; WX, 82270938), CAMS Innovation Fund for Medical Sciences (WX, 2021-I2M-1-002), National Key R&D Program of China (WX, 2021YFC2501700), National High Level Hospital Clinical Research Funding (WX, 2022-PUMCH-D-006), the Non-profit Central Research Institute Fund of Chinese Academy of Medical Sciences (LC, 2023-PT320-10), and Young Elite Scientists Sponsorship Program by BAST (LC, No.BYESS2023171). Part of the study was supported by Merck Sharp & Dohme China, Hangzhou, China.

**Research in context:** *Evidence before this study:* Bone mineral density (BMD) from dual X-ray absorptiometry was firstly used to predict fragility fracture, but had low sensitivity. Tools like FRAX, QFracture, and Garvan, which also incorporated clinical factors into prediction models, showed improved performance. Models containing standard HR-pQCT indices (μFRAC) further surpassed most clinical tools. Nevertheless, direct learning from original HR-pQCT images is always desired to reduce labor and bias. Deep learning being the most common method for image-based learning, we searched PubMed for articles published up to Mar 25, 2024, using keywords “(‘fragility fracture’ OR ‘osteoporotic fracture’) and (‘prediction model’) and (‘HR-pQCT’ or ‘High-resolution peripheral quantitative CT’) and (‘deep-learning’ OR ‘deep learning’)”. Results showed that no study has built deep learning models from HR-pQCT for fragility fracture prediction.

*Added value of this study:* We developed DeepQCT from HR-pQCT of 216 elderly Chinese women from a national cohort (ChiVOS), which calculated risk scores using individual bone images and clinical features. BMD and pQCT-index models were compared to DeepQCT. We found both DeepQCT (best model AUC 0.86-0.94) and pQCT-index (best model AUC 0.8-0.93) outperformed BMD (best model AUC 0.54-0.78). DeepQCT using non-weight-bearing bones (ulna, fibula) performed similarly to weight-bearing bones (tibia, radius). Specifically, HR-pQCT revealed one patient subgroup with higher DeepQCT risk scores, which showed lower BMD and multiple bone microarchitectural abnormalities, associated with osteoporosis and fragility fracture history.

*Implications of all the available evidence:* DeepQCT is the first method which uses deep-learning to predict fragility fracture directly from HR-pQCT images. It is also the first to use single bones individually in prediction models, including non-weight-bearing bones, which are excluded in HR-pQCT-index computation. Of note, DeepQCT risk score is highly clinically relevant, as showed in bone density or microarchitectural features differences between patient subgroups. The non-inferior performance of DeepQCT compared to the manual annotation-dependent pQCT-index, supported its application to reduce labor and enhance efficiency. Performance of non-weight-bearing bones also challenges traditional perception of using load-bearing bones only in predicting osteoporotic conditions. Most importantly, the DeepQCT framework can be easily adapted for other tasks using HR-pQCT scans, which greatly expands application of digital medicine in bone mineral disease diagnosis or management.

## Introduction

Osteoporosis is a disease of bone mass decrease and microstructure deterioration, leading to increased fragility fracture risk. Osteoporosis is highly prevalent in elderly postmenopausal women. In 2017-2018, its prevalence was 19.6% in American women aged 50 years or older^1^, and 20.6% in Chinese women aged 40 years or older^2^, with estimated fragility fractures reaching 4.3 million in Europe^3^, and 2.3 million in US^4^. Osteoporotic fractures mainly happen in hip, vertebrae, and wrist, which cause increased mortality, morbidity, and economic burden. E.g., mortality ratio of hip fracture was around 2-fold^5^ than those without fracture, and osteoporotic fracture treatment annually costed over 5 trillion USD in Europe and North America^6^. Of note, osteoporosis is underdiagnosed and undertreated^7^, which made primary prevention essential^8^. Importantly, future fracture screening has proved beneficial and cost-effective in elderly European women from randomized controlled trials^9,10^.

Various tools were developed to assess bone fragility. In 1994, WHO designated diagnostic criteria of osteoporosis as having dual x-ray absorptiometry (DXA)-based areal bone mineral density (aBMD) T score of -2.5 or lower, which remained the golden standard today. However, areal BMD was computed from 2-dimensional images, which differed from the real volumetric BMD. In fact, the BMD method suffered from low sensitivity around 30 to 50% in predicting fracture^11^. This prompted development of alternative prediction tools – e.g., the fracture risk assessment (FRAX) tool. Developed in 9 and validated in 11 cohorts worldwide, the FRAX tool estimated major osteoporotic fracture probability in 10-years, using femoral neck BMD, body mass index, fracture history, parental hip fracture history, tobacco smoking, glucocorticoid usage, rheumatoid arthritis, and alcohol consumption^12,13^. Similarly, QFracture and Garvan were also consisted of clinical risk factors, and were developed and validated in large cohorts, from the United Kingdom and Australia respectively^14,15^.

Despite great usability, clinical factors do not directly reflect bone structures. Beyond DXA, research tools were developed for refined profiling of bone microstructures, with one highlight being the quantitative CT (QCT). QCT, either central or peripheral, produces multiple 2D slices, thus allowing for calculation of volumetric bone density (vBMD) at vertebrae or limb bones (radius and tibia). The high-resolution peripheral QCT (HR-pQCT) was later developed^16^, which permits semi-automated computation of multiple geometric, microarchitectural, and biomedical features of bones, other than vBMD^17,18^. Currently, with the second-generation scanner becoming available, HR-pQCT has been applied in multiple research fields and in some clinical scenarios^19^.

Many studies have since investigated associations of HR-pQCT indices with osteoporotic fractures. Of note, HR-pQCT indices showed significant difference in patients with and without a history of fragility fractures^20-23^, and were related to future incident fractures^16,24^. Remarkably, HR-pQCT indices could predict fracture risk independently from femoral neck aBMD or FRAX score; and cortical density, trabecular number, and trabecular thickness were reported as best predictors in both distal radius and tibia^25^. One step further, the μFRAC tool, a random forest classifier, used HR-pQCT indices to predict osteoporotic fracture, and outperformed FRAX and femoral neck aBMD^26^. Likewise, models using texture analysis from original HR-pQCT images also showed improved performance in discriminating fracture history than BMD and clinical covariates^27^. However, HR-pQCT used by most studies were computed by scanner based on manual annotation on radius or tibia, which could cause bias by excluding non-weight-bearing bones (fibula and ulna), and inaccuracy from technical variance among annotators. Similar drawbacks lie in radiomics (e.g. texture analysis), as quantification by pre-defined metrics risks loss of other features. Direct learning from image is thus desired. The deep learning (DL) method based on neural network is a perfect match for this goal. Using images directly as input, DL has showed supreme diagnostic performance in pathology^28,29^, dermatology^30,31^, and radiology^32,33^. Specifically, it was also able to diagnose rheumatoid and psoriatic arthritis from hand HR-pQCT^34,35^, making it a promising solution also for limb HR-pQCT.

We here aim to construct a deep learning model, DeepQCT, to predict fragility fracture history from HR-pQCT images. Specifically, models of individual bones will be assessed, along with HR-pQCT-index or BMD models. Furthermore, we aim to interpret DeepQCT using bone density or microarchitectural phenotypes in patient subgroups.

## Materials and Methods

### HR-pQCT ChiVOS cohort and study design

Recruitment of the Chinese Vertebral Osteoporosis Study (ChiVOS) cohort was previously described.^36,37^ In brief, ChiVOS was a national community-based cohort of 2,664 postmenopausal women aged 50 or older in China. All participants answered survey questionnaires and received physical exams to collect age, height, weight, and history of fragility fractures. They also received dual x-ray absorptiometry (DXA) and lateral thoracic and lumbar spine X-rays. Specifically, fragility fracture history was positive if patient answered ‘yes’ in questionnaire for ‘ever having fracture from low energy trauma such as fall from standing height after age of 50’, or if vertebral fracture was identified from lateral X-ray of thoracic or lumbar spine using Genant’s semiquantitative methods^38^. HR-pQCT scans were performed at non-dominant distal arm or leg in patients who agreed scanning at Peking Union Medical College Hospital (PUMCH). The final HR-pQCT ChiVOS cohort included only patients with HR-pQCT images at both distal radius and tibia (N=267), and excluded those with missing DXA data at either femoral neck, total hip, or lumbar spine (L1-L4), resulting in 216 patients. For model training, 21 patients (10%) were randomly selected as internal test set, and the rest 195 patients (90%) were used as development set. Within the development set, six-fold cross-validation was performed, generating 162 patients in training set and 33 in validation set at each fold.

### DXA and HR-pQCT scans

DXA scans (GE-Lunar scanners, GE Healthcare, Madison, WI, USA) were performed in all participants at femoral neck, total hip, and lumbar spine (L1-L4) by certified technicians at PUMCH. Areal bone mineral density (aBMD) was measured at each site (FN BMD, TH BMD, LS BMD). Osteoporosis by aBMD (osteoporosis) was positive if patient had T score ≤ -2.5 in femoral neck, total hip, or L1-L4, regardless of fragility fracture history. HR-pQCT scanning (Xtreme CT II; Scanco Medical AG, Bassersdorf, Switzerland) was performed at distal radius (1^st^ slice at 9.0mm proximal to reference lines) and tibia (1^st^ slice at 22.0mm proximal to reference lines) at nondominant side of patients using standard protocol^39^, which generated 168 CT slices of 2304 x 2304 pixels at 61uM resolution (x and y axis) within range of 1cm (z axis). For each scan, trained technicians annotated key structures on radius or tibia, from which standard HR-pQCT indices were calculated by scanner (Xtreme CT II, 3D Density and Structure Analysis). These indices included total bone area (Tt.Ar), trabecular area (Tb.Ar), cortical area (Ct.Ar), cortical perimeter (Ct.Pm), total bone volumetric BMD (Tt.vBMD), trabecular volumetric BMD (Tb.vBMD), cortical volumetric BMD (Ct.vBMD), bone volume fraction (BV/ TV), trabecular number (Tb.N), trabecular thickness (Tb.Th), trabecular separation (Tb.Sp), cortical thickness (Ct.Th), and cortical porosity (Ct.Po).

### Image preprocessing of HR-pQCT scans

To prepare for DeepQCT training, we segmented original HR-pQCT scans to images of individual bones (radius and ulna for upper limb scans, and tibia and fibula for lower limb scans). In brief, 116 patients were randomly selected from 2,898 patients of in-house PUMCH cohort who had HR-pQCT scans of both distal radius and tibia (same protocol as HR-pQCT in ChiVOS), from which bone contours were manually labeled on scans (ITK-SNAP, v3.8.0). A U-Net was trained with 10 times augmentation with default parameters (MONAI, v1.3.0), which was then applied to all scans in HR-pQCT ChiVOS to calculate contour masks of bones. Using these masks, we computed horizontal and vertical margins of each bone by extracting connected regions (skimage, v0.19.3), which eventually generated cropped images of radius, ulna, tibia, and fibula for each participant. Manual examination of the images confirmed correct and clean segmentation in nearly all cases.

### DeepQCT models

For DeepQCT, we first employed ResNet34 (Medical Open Network for AI, MONAI^40^) to generate a 128-dimensional output vector (DeepQCT vector). Specifically, we developed sub-models with either one input channel of single bones (radius, ulna, tibia, or fibula), or four input channels of all bones (radius, ulna, tibia, and fibula). Multi-task learning was used in training, which incorporated age, sex, weight, height, and HR-pQCT indices from both radius and tibia into loss function by cross-entropy (cross-entropy) for categorical variables, and mean squared error (MSE) for numeric variables (Loss = Cross Entropy_fracture_ + 0.01 × Cross Entropy_sex_ + 0.01 × MSE_age,weight,height_ + 0.1 × MSE_HR-pQCT indicates (radius and tibia)_). Each model was trained with 4-time augmentation at learning rate of 0.001 with Adam optimizer. In each fold, the best model was chosen based on the maximum Area Under the ROC Curve (AUC) score from the validation set, employing early stopping after 200 epochs.

The final DeepQCT models were sur-named with ulna, radius, fibular, tibia or all bones, based on which bone image were used in the ResNet step. Final AUC scores were computed in test set using logistic regression of DeepQCT vector and clinical factors (age, sex, height, weight) at each fold. For each model, the optimal cut-off was determined by maximizing the difference between the True Positive Rate (TPR) and the False Positive Rate (FPR).

### Other predictive models

Multivariant logistic regression was used to construct other models with the same training, validation, and testing dataset as DeepQCT. Specifically, the BMD model included age, sex, height, weight, and BMDs (FN BMD, TH BMD, and LS BMD). The pQCT-index models included age, sex, height, weight, and standard HR-pQCT indices of radius (for upper limb), tibia (for lower limb), or both radius and tibia (all-bone model). AUCs and optimal cut-off threshold were determined with the same methods as DeepQCT.

### Other statistical analysis

Comparisons of demographic features and HR-pQCT indices between patients with or without history of fragility fracture were conducted using Mann-Whitney U test for continuous variables, and Chi-squared test for categorical variables. Spearman’s rank correlation was used to calculate pairwise correlation of HR-pQCT indices among patients. Patient clustering was performed using cosine similarity and average linkage from Spearman’s rank correlation matrix using HR-pQCT indices. The two patient clusters were obtained at the first hierarchy. For dimensionality reduction, principal component analysis (PCA) was performed using DeepQCT vector. Comparisons of HR-pQCT indices between patient subgroups were performed using Mann-Whitney U test with Benjamini-Hochberg adjustment.

## Results

### Patient characteristics

The HR-pQCT ChiVOS cohort consisted of 216 postmenopausal Chinese women at or over 50 years with complete data of demographic features, aBMDs, and HR-pQCT scans (Fig. 1, Methods), which included 62 individuals (28.7%) with fragility fracture history (FF group), and 154 (71.3%) without fragility fracture history (Ref group). In general, patients in FF group were older, slightly shorter, and had lower femur aBMD. However, LS BMD or TH BMD of both groups were not significantly different. In radius, FF group had smaller cortical area (48 vs 52mm^2^), fewer trabecular number (0.99 vs 1.1mm^-1^), higher trabecular separation (1.2 vs 0.95mm), and lower cortical thickness (0.88 vs 0.95mm), as well as lower total vBMD (240 vs 270mg/cm^2^), cortical vBMD (870 vs 890mg/cm^2^), and trabecular vBMD (85 vs 100mg/cm^2^) than Ref. In tibia, FF group had smaller cortical area (91 vs 100mm^2^), lower cortical thickness (1.1 vs 1.2mm), lower total vBMD (210 vs 230mg/cm^3^), and lower cortical vBMD (800 vs 840mg/cm^3^). Of note, most FF group were also osteoporotic (77.4%), while most patients of Ref group had no osteoporosis (83.8%) (Table 1).

**Table 1.** Demographic features, aBMD T scores, and HR-pQCT indices in patients with and without fragility fracture history in HR-pQCT ChiVOS cohort.

| Group | No fragility fracture history <sup>a</sup> | With fragility fracture history | P-value <sup>b</sup> |
| --- | --- | --- | --- |
| Number (%) | 154 (71.3) | 62 (28.7) | / |
| Age |  |  |  |
| ≥50, <60 | 41 (26.6%) | 5 (8.1%) | <0.001 |
| ≥60, <70 | 66 (42.9%) | 25 (40.3%) |  |
| ≥70, <80 | 37 (24.0%) | 19 (30.6%) |  |
| ≥80 | 10 (6.5%) | 13 (21.0%) |  |
| Height (m) | 1.6 ± 0.061 | 1.5 ± 0.067 | 0.0162 |
| Weight (kg) | 62 ± 11 | 62 ± 9.8 | 0.996 |
| Osteoporosis by aBMD <sup>c</sup> |  |  |  |
| No | 129 (83.8%) | 14 (22.6%) | <0.001 |
| Yes | 25 (16.2%) | 48 (77.4%) |  |
| aBMD T score |  |  |  |
| Femoral neck |  |  |  |
| ≥-1 | 66 (42.9%) | 19 (30.6%) | 0.00461 |
| <-1, >-2.5 | 74 (48.1%) | 27 (43.5%) |  |
| ≤-2.5 | 14 (9.1%) | 16 (25.8%) |  |
| Lumbar spine (L1-L4) |  |  |  |
| ≥-1 | 87 (56.5%) | 35 (56.5%) | 0.435 |
| <-1, >-2.5 | 51 (33.1%) | 17 (27.4%) |  |
| ≤-2.5 | 16 (10.4%) | 10 (16.1%) |  |
| Total hip |  |  |  |
| ≥-1 | 83 (53.9%) | 25 (40.3%) | 0.13 |
| <-1, >-2.5 | 62 (40.3%) | 30 (48.4%) |  |
| ≤-2.5 | 9 (5.8%) | 7 (11.3%) |  |
| HR-pQCT indices (Radius) <sup>d</sup> |  |  |  |
| Geometry (mm<sup>2</sup>) |  |  |  |
| Tt.Ar | 250 ± 39 | 250 ± 40 | 0.289 |
| Ct.Ar | 52 ± 10 | 48 ± 9.4 | 0.00794 |
| Tb.Ar | 200 ± 40 | 200 ± 42 | 0.632 |
| Ct.Pm | 66 ± 6.2 | 65 ± 5.6 | 0.602 |
| Microarchitecture |  |  |  |
| BV/TV | 0.15 ± 0.052 | 0.14 ± 0.041 | 0.0517 |
| Tb.N (mm<sup>-1</sup>) | 1.1 ± 0.29 | 0.99 ± 0.31 | 0.00218 |
| Tb.Th (mm) | 0.22 ± 0.017 | 0.22 ± 0.029 | 0.414 |
| Tb.Sp (mm) | 0.95 ± 0.36 | 1.2 ± 0.56 | 0.002 |
| Ct.Th (mm) | 0.95 ± 0.22 | 0.88 ± 0.21 | 0.045 |
| Ct.Po (%) | 0.0073 ± 0.0047 | 0.0090 ± 0.0060 | 0.0619 |
| **Volumetric BMD (mg/cm<sup>3</sup>)** |  |  |  |
| Tt.vBMD | 270 ± 79 | 240 ± 69 | 0.0375 |
| Ct.vBMD | 890 ± 72 | 870 ± 71 | 0.0331 |
| Tb.vBMD | 100 ± 40 | 85 ± 36 | 0.0141 |
| **HR-pQCT indices (Tibia)** |  |  |  |
| **Geometry (mm<sup>2</sup>)** |  |  |  |
| Tt.Ar | 660 ± 98 | 660 ± 89 | 0.607 |
| Ct.Ar | 100 ± 21 | 91 ± 18 | 0.00505 |
| Tb.Ar | 560 ± 100 | 570 ± 91 | 0.333 |
| Ct.Pm | 99 ± 7.6 | 100 ± 6.9 | 0.272 |
| **Microarchitecture** |  |  |  |
| BV/TV | 0.19 ± 0.046 | 0.18 ± 0.042 | 0.24 |
| Tb.N (mm<sup>-1</sup>) | 1.1 ± 0.22 | 1.0 ± 0.27 | 0.154 |
| Tb.Th (mm) | 0.24 ± 0.020 | 0.25 ± 0.023 | 0.082 |
| Tb.Sp (mm) | 0.94 ± 0.25 | 1.1 ± 0.48 | 0.0882 |
| Ct.Th (mm) | 1.2 ± 0.27 | 1.1 ± 0.22 | 0.014 |
| Ct.Po (%) | 0.035 ± 0.015 | 0.038 ± 0.016 | 0.0974 |
| **Volumetric BMD (mg/cm<sup>3</sup>)** |  |  |  |
| Tt.vBMD | 230 ± 61 | 210 ± 44 | 0.00603 |
| Ct.vBMD | 840 ± 69 | 800 ± 64 | <0.001 |
| Tb.vBMD | 120 ± 37 | 110 ± 35 | 0.216 |

<sup>a</sup> Continuous variables shown as mean ± standard deviation; categorical variables shown as frequency and percentage.
<sup>b</sup> Mann-Whitney U test for continuous variables; Chi-squared test for categorical variables.
<sup>c</sup> Osteoporosis by aBMD defined by T score ≤ -2.5 in femoral neck, total hip, or lumbar spine (L1-L4).
<sup>d</sup> Tt.Ar, total bone area; Tb.Ar, trabecular area; Ct.Ar, cortical area; Ct.Pm, cortical perimeter; Tt.vBMD, total bone volumetric BMD; Tb.vBMD, trabecular volumetric BMD; Ct.vBMD, cortical volumetric BMD; BV/TV, bone volume fraction; Tb.N, trabecular number; Tb.Th, trabecular thickness; Tb.Sp, trabecular separation; Ct.Th, cortical thickness; Ct.Po, cortical porosity.

**Figure 1.**
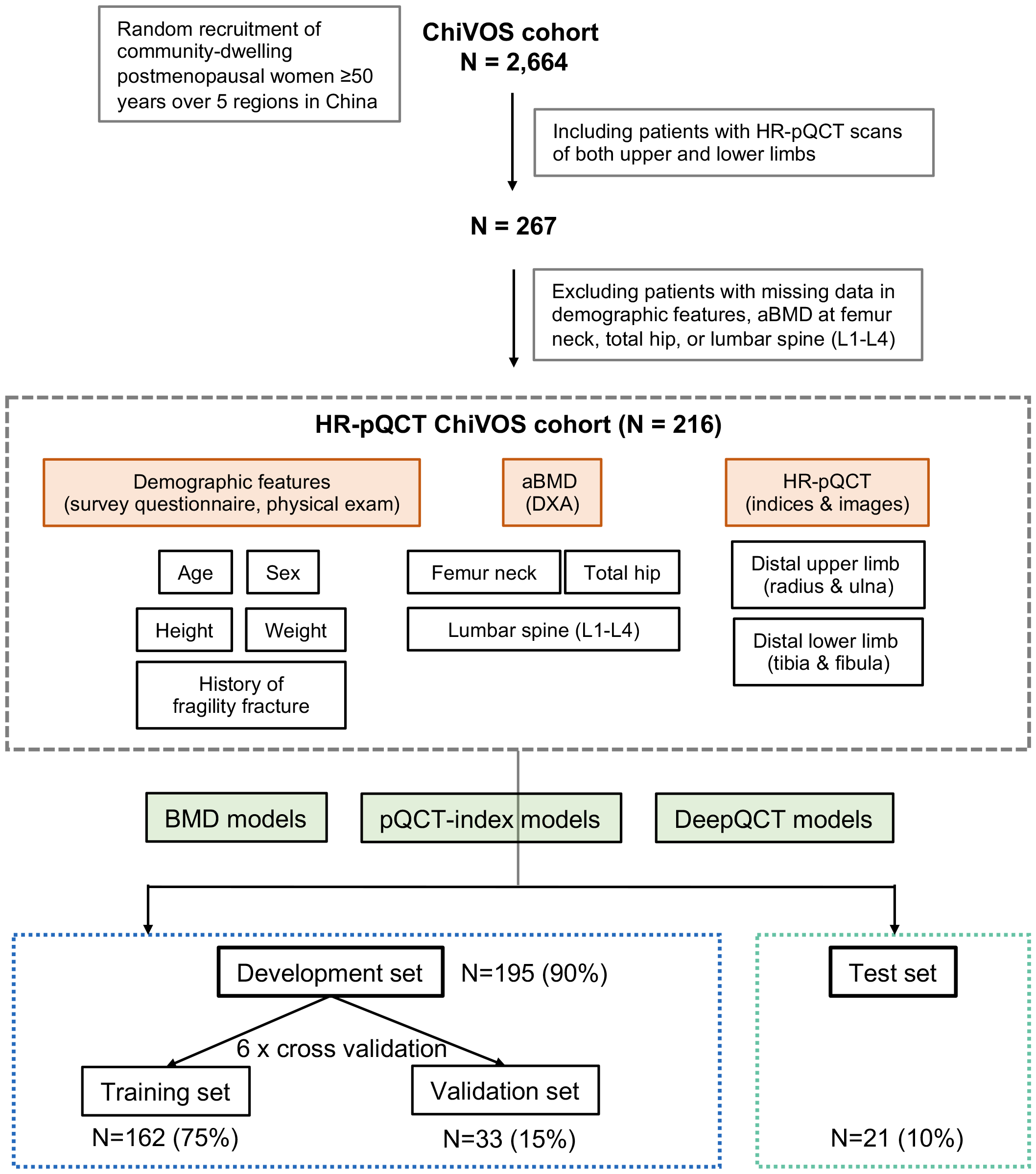
HR-pQCT ChiVOS cohort and study design. Establishment of HR-pQCT ChiVOS cohort and workflow of model assessment. Inclusion and exclusion criteria, prediction models types, and data splitting for model training and testing were illustrated.

### DeepQCT and pQCT-index models outperform BMD models

We constructed three types of models to predict fragility fracture history. To increase interpretability, logistic regression was used as last step for all models. The BMD models most resemble current clinical practice, which used age, height, weight, FN BMD, LS BMD, and TH BMD in logistic regression. The pQCT-index model also used age, height, weight, and additionally standard HR-pQCT indices. Three sub-models were included, which used indices of radius, tibia, or both (radius+tibia) respectively. The DeepQCT models used HR-pQCT images directly and included five sub-models, using scans of radius, ulna, tibia, or fibula as single channel input, or all four bones as four-channel input respectively (Fig. 2A). Given minimal variation observed in scans along z-axis, one single image was randomly selected from the 168 scans, as input of DeepQCT. To improve performance, we used multi-task learning to prompt concomitant prediction of age, sex, and HR-pQCT indices from both radius and tibia of the same patient. The output from ResNet, a 128-dimensional vector, was subsequently merged with age, sex, weight, and height in the final logistic regression, to predict a numeric probability and a binary label. Computation was performed for each model at each cross-validation fold.

**Figure 2.**
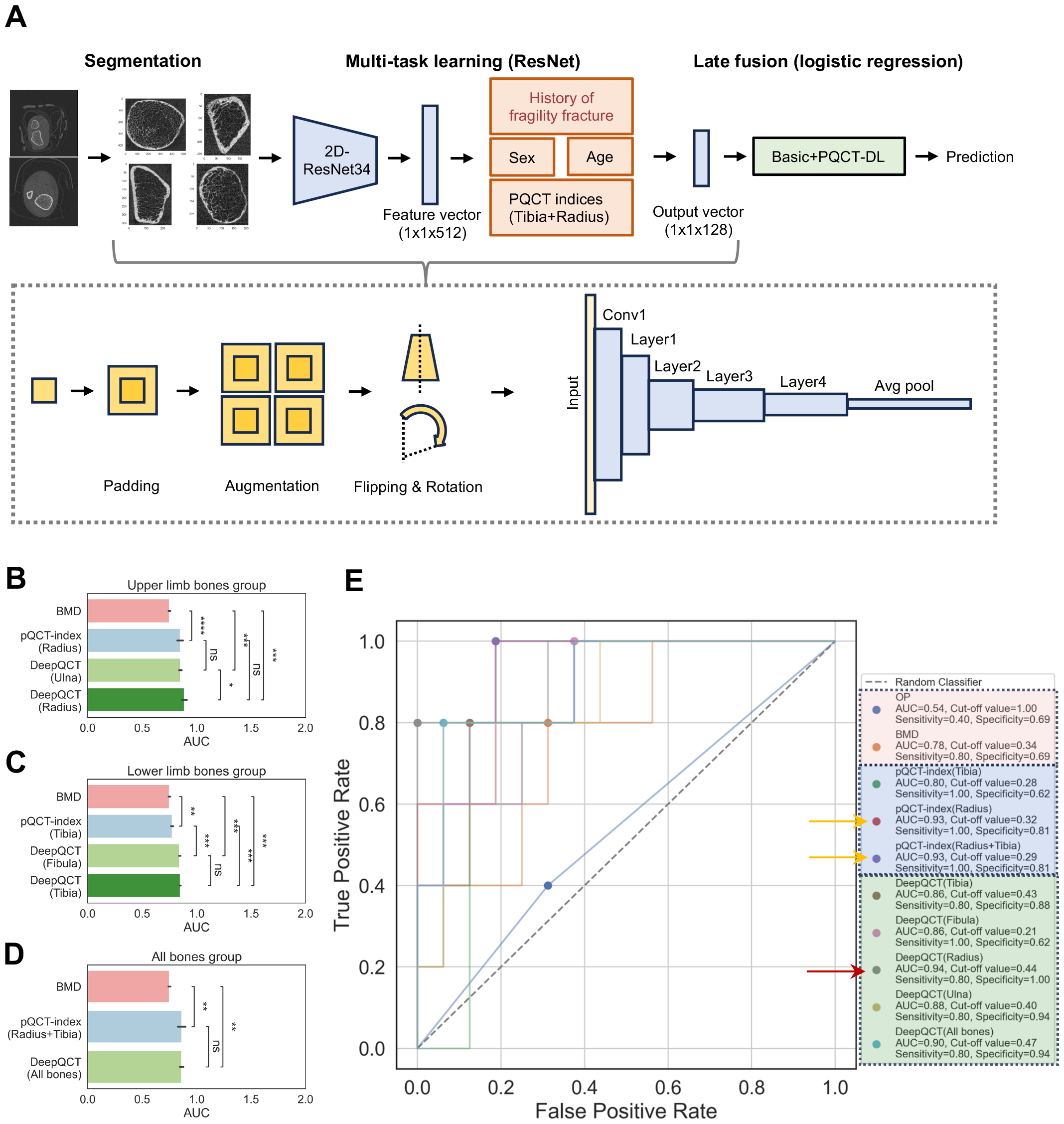
Construction and performance of all models predicting fragility fracture history. A. Workflow of image preprocessing, and DeepQCT construction. B-D. AUC scores of all sub-models in groups of the upper limb bones (B), lower limb bones (C), or all bones (D). Error bars showing variation from cross-validation, with significance by Mann-Whitney U test (*0.01<p≤0.05, **0.001<p≤0.01, ***0.0001<p≤0.001, ns, p>0.05). E. Receiver operator curve of best-performing models from cross validation, showing AUC, optimal cut-off value of predicted probability, and sensitivity and specificity at optimal cut-off. Pink box, OP (osteoporosis) or BMD models; blue box, pQCT-index models; green box, DeepQCT models. Red arrow, DL-radius model with top AUC (0.94). Yellow arrows, pQCT-index models with second-top AUC (0.93) of radius, and radius+tibia.

We assessed model performance using area under the curve (AUC). Specifically, pQCT-index or DeepQCT models were compared within three groups across all cross-validation folds – those using upper limb bones, lower limb bones, or all bones. BMD models were included in all groups as reference. In the upper limb bones group, both DeepQCT (ulna and radius) and pQCT-index models significantly outperformed BMD, with radius-DeepQCT outperforming ulna-DeepQCT. DeepQCT had higher average AUCs than pQCT-index model, but with no significant difference (Fig. 2B). In the lower limb bones group, DeepQCT and pQCT-index models were both superior than BMD, and both fibula-DeepQCT and tibia-DeepQCT outperformed pQCT-index models (Fig. 2C). In the all-bones group, DeepQCT and pQCT-index performed similarly, but both surpassed BMD models (Fig. 2D). Overall, HR-pQCT data significantly improved fragility fracture prediction than areal bone mineral density. In lower limbs, original HR-pQCT images further improved model performance than pQCT-index. Interestingly, radius-DeepQCT only slightly outperformed that of ulna, and tibia and fibula-DeepQCT showed similar performance. This suggested that ulna and fibula hold important osteoporotic characteristics as weight-bearing bones, and should not be empirically excluded from fragility fracture analysis.

Receiver operating characteristic (ROC) curves were also assessed to obtain optimized threshold of prediction score from each model (Fig. S1). Specifically, the best-performing one from 6 cross-validation was selected for each sub-model type (Fig. 2E). An osteoporosis diagnosis model based solely on aBMD was incorporated (OP model), wherein a positive prediction was defined as having a T score of -2.5 or below in one or more of the FH, TH, or LS BMD. From AUC, all models outperformed the OP model, among which all DeepQCT or pQCT-index models surpassed the BMD model. Specifically, radius-DeepQCT had highest AUC of 0.94, with sensitivity of 0.8 and specificity of 1 using cut-off value at 0.44 (red arrow, Fig. 2E). This was closely followed by pQCT-index model of radius (AUC 0.93, sensitivity 1 and specificity 0.81 with cut-off value at 0.32), and pQCT-index model of tibia and radius (AUC 0.93, sensitivity 1 and specificity 0.81 with cut-off value of 0.29) (yellow arrows, Fig. 2E). Interestingly, for each best-performing pQCT-index or DeepQCT model, optimal cut-off of risk score was smaller than 0.5, which suggested raising awareness even with low predicted probability.

### HR-pQCT indices correlate with one another and with fragility fracture

Next, we investigated correlation between different HR-pQCT indices, and their association with fragility fracture history. Hierarchical clustering was performed using pairwise correlation between indices, which were annotated with source bone (radius or tibia) and parameter groups, including geometric (Tt.Ar, Ct.Ar, Tb.Ar, Ct.Pm), microarchitectural (BV/TV, Tb.N, Tb.Th, Tb.Sp, Ct.Th, Ct.Po), and vBMD (Tt.vBMD, Ct.vBMD, Tb.vBMD) (Fig. 3A).

**Figure 3.**
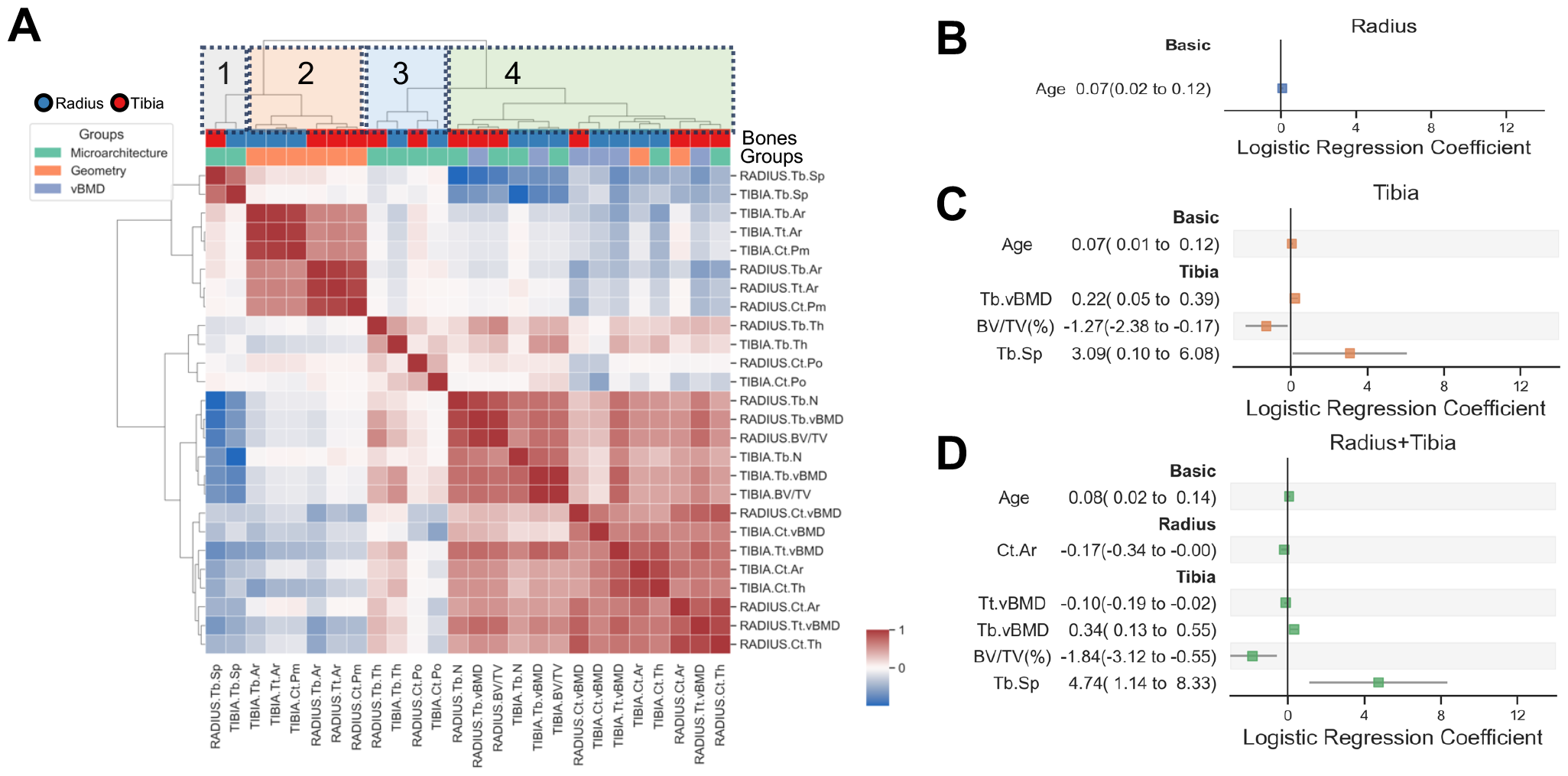
Correlation of HR-pQCT indices and logistic regression of pQCT-index models. A. Spearman’s rank correlation of HR-pQCT indices of radius and tibia, showing source bone and index groups on top bars, and four clusters in top boxes. B-D. Logistic regression coefficient of each model type with 95% confidence interval from pQCT-index models of radius (B), tibia (C), and radius+tibia (D). Only predictors with significance (p<0.05) were shown.

Four major clusters were derived. The first contained trabecular separation of both tibia and radius, and cluster 2 or 3 contained most other microarchitecture components. Specifically, cluster 2 comprised almost all geometric indices, including perimeter and area size, while cluster 3 comprised trabecular thickness and cortical porosity. Interestingly, vBMD (total, cortical, and trabecular), which was intrinsically associated with osteoporosis, were all located in cluster 4. Trabecular number and bone volume fraction (BV/TV) were also in cluster 4. Specifically, cortical area size or thickness were positively related to vBMD within the same cluster (cluster 4), in contrast to trabecular area size or thickness, which were less relevant. This showed HR-pQCT clustering were only partially affected by index or bone types, and that certain microarchitectural features were more related to volumetric BMDs than others.

We next examined association of HR-pQCT indices to fragility fracture history from the three pQCT-index models of logistic regression (Fig. 3B-D). As expected, age was a significant risk factor of fragility fracture history (FFH) in all models. In both tibia and radius+tibia models, tibial bone volume fraction (BV/TV%) was consistently protective, whereas tibial trabecular separation was consistently a risk factor. Both total vBMD and radial cortical area were also protective in radius+tibia model. Unexpectedly, tibia trabecular vBMD was found positively correlated with FFH, opposite from total vBMD. While this could result from erroneous inclusion of cortical in trabecular areas by machine due to vague separation in osteoporosis, it indicated necessity to assess total, cortical, and trabecular vBMD, rather than trabecular vBMD alone. In summary, pQCT-index models suggested that older age, higher tibia trabecular separation, lower tibial bone volume fraction, lower radial cortical area, and lower tibial total vBMD correlated with increased risk of fragility fracture.

### Identification of a patient subgroup susceptible to fragility fractures

We then aim to classify patients by bone density and microstructural features from HR-pQCT. From correlation matrix of HR-pQCT indices, patients formed two distinct subgroups (Fig. 4A). Interestingly, subgroup 1 showed higher ratio in both fragility facture history and osteoporosis by aBMD (Fig. 4B). Consistently, these two subgroups were separated in the 3-dimensional principal component space derived from the DeepQCT vector (Fig. 4C), which suggested intrinsic distinct HR-pQCT phenotypes in bone images.

**Figure 4.**
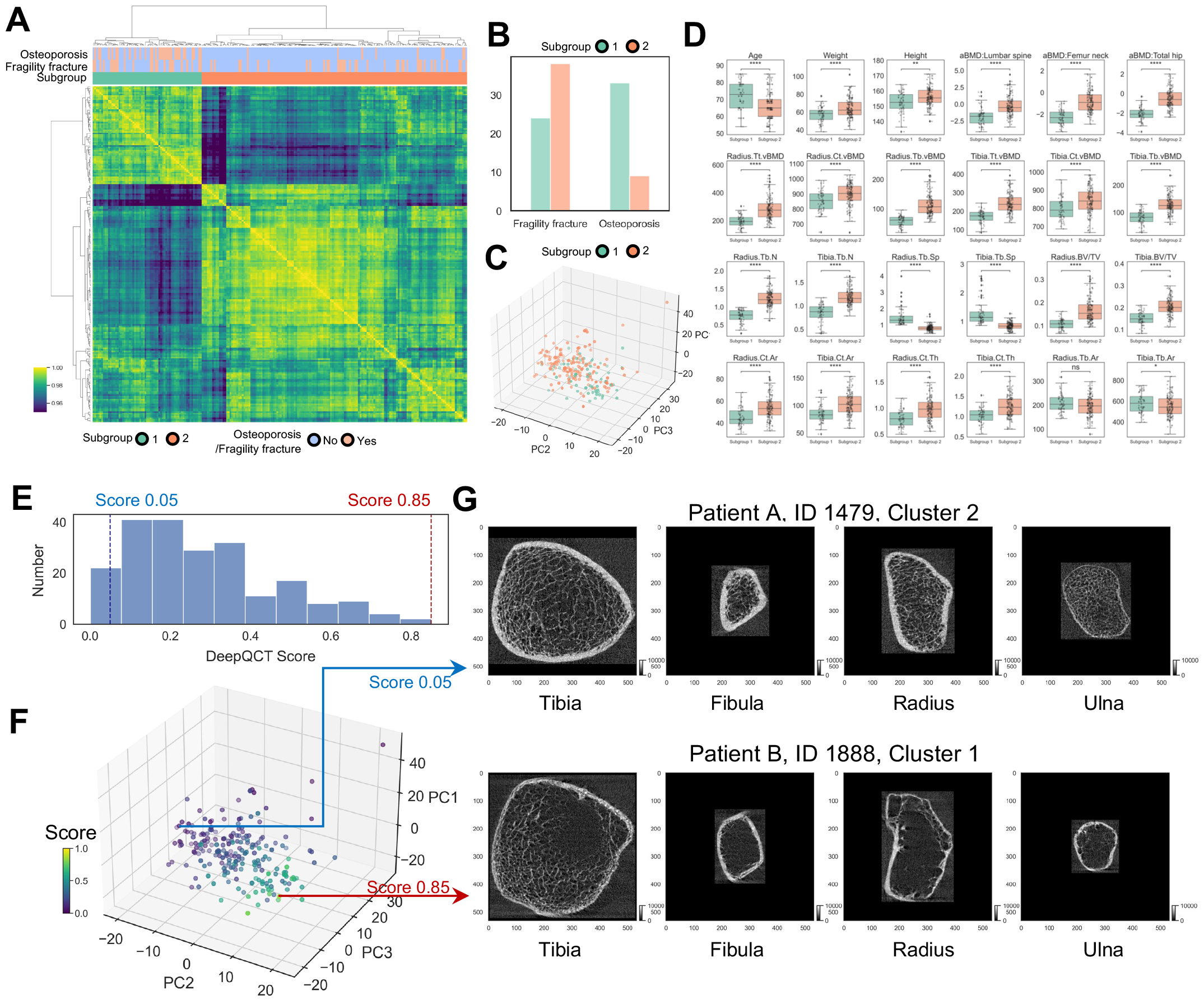
Identification and characteristics of patient subgroups using HR-pQCT. A. Patient clustering using Spearman’s rank correlation matrix of HR-pQCT indices from both radius and tibia. B. Ratio of having fragility fracture history and osteoporosis by aBMD in patient subgroups. C. Scatter plot of patient subgroups in the top three principal component space derived from DeepQCT vector. D. Distribution of demographics, aBMD, and HR-pQCT indices in patient subgroups with significant difference between two groups in either radius or tibia (Mann-Whitney U test, *0.01<p≤0.05, **0.001<p≤0.01, ***0.0001<p≤0.001, ns, p>0.05). E. Histogram of DeepQCT score representing predicted probability of fragility fracture history. F. DeepQCT scores of patients in the top three principal component space derived from DeepQCT vector. G. HR-pQCT images of tibia, fibula, radius, and ulna in representative patients. Scores and PCA embeddings all based on all-bones DeepQCT model fold 4, as illustration representative.

Next, we examined differences between two patient subgroups (Fig. 4D). In general, patients of subgroup 1 were older, lighter, and shorter. Of note, all BMD indices were lower in cluster 1, including aBMD at femoral neck, lumbar spine, and total hip, as well as total, cortical, and trabecular vBMD at radius and tibia. Additionally, subgroup 1 also had fewer trabecular number, higher trabecular separation, lower bone volume fraction, smaller cortical area, and lower cortical thickness in both radius and tibia (Fig. 4D). Trabecular area was larger in subgroup 1 than 2 for tibia, while similar in both groups for radius. Of note, besides vBMD, most features significantly different in two clusters was microarchitecture, rather than geometry. In summary, subgroup 1 patients had significant aberrant bone microarchitecture and lower bone mineral density, which likely explained their higher vulnerability to fragility fracture.

Finally, we integrated DeepQCT with clinical cases. Overall, DeepQCT risk scores were mono-modal and skewed to the left (Fig. 4E). In 3-dimensional PCA space, patients with higher DeepQCT risk scores overlap more with patient subgroup 1, which was more vulnerable to fragility fracture (Fig. 4C, F). Two patients with either low or high DeepQCT risk scores were demonstrated as representatives - patient A from cluster 2 with DeepQCT score of 0.04, and patient B from cluster 1 with DeepQCT score of 0.84. From original HR-pQCT scans, bone size was generally similar between two patients in tibia, fibula, and radius, though patient B had a smaller ulna, likely as a common biological variant. Specifically, patient A exhibited higher bone density (higher image brightness), thicker cortical bone, and well-defined, uniform trabecular structures. In contrast, patient B showed lower bone density (lower image brightness), thinner cortical layer, and less distinct, more sparse trabecular bone patterns (Fig. 4G). Consistently, clinical information of patient B revealed a lady of 80-85 years with osteoporosis, who experienced a vertebral fracture, a distal radial fracture, and a proximal humoral fracture during 1991 to 1996, and a knee fracture during 2010-2015. Patient A, conversely, was a lady of 50-55 years with neither osteoporosis nor history of any form of fracture. These examples verified significant clinical and pathological difference in patients with distinct predicted risk by DeepQCT model.

## Discussion

In this study, we created DeepQCT, which predicts fragility fracture history directly from HR-pQCT images in elderly Chinese women. Impressively, DeepQCT outperformed BMD models, and surpassed pQCT-indices in selected conditions. Furthermore, large DeepQCT scores were highly consistent with a patient subgroup with low volumetric BMD and multiple bone microarchitectural abnormalities, who are more susceptible to osteoporosis or fragility fracture. These abnormalities are clearly visualized from original scans at distal limbs.

DeepQCT is the first automatic pipeline which uses HR-pQCT scans directly to predict fragility fracture history. This included two-steps – first, we trained a segmentation model (U-Net) which detect bone contours; second, we built a classification model (ResNet) which permits fusion with non-image factors. Our ResNet model was optimized in performance and flexibility. First, image augmentation expanded training set to four times with random flipping and rotation, to promote robust training. Second, we set side task for the model to learn fragility-fracture-relevant factors, including age, height, weight, and HR-pQCT indices, to further enhance performance. Third, the late fusion step created high flexibility for both non-image variable inclusion and algorithm selection. From performance by AUC, DeepQCT was superior to BMD models. While only being non-inferior to pQCT-index models, DeepQCT was a fully-automatic, which spared the time-consuming and error-prone manual labeling in pQCT-index models, especially with malformed bone structures. Moreover, with such flexibility, DeepQCT can be easily adapted for other diagnostic tasks using HR-pQCT scans. For example, it can be trained to discriminate multiple osteoporotic conditions of X-linked hypophosphatemic rickets, tumor-induced osteomalacia, and primary hyperparathyroidism, showcasing wide utility in advancing bone health with digital medicine.

Another highlight is that we assessed predictive value of individual bones, by segmenting the whole limb scan to bone-specific images. Traditionally, only weight-bearing bones were included in either standard HR-pQCT index computation, or in prediction models. Ulna or fibula were thus seldomly used. In our analysis, DeepQCT models of ulna or fibula were similar or only slightly inferior to their weight-bearing counterparts (radius or tibia). This supported including non-weight bearing bones in osteoporotic fracture prediction and bone microarchitectural analyses.

Detailed assessment of HR-pQCT indices highlighted specific parameters likely more important to fragility fracture. From hierarchical clustering, we noticed that only a subset of microstructural indices closely and positively correlated to vBMD - these included trabecular number, bone volume fraction (BV/TV), cortical area size, and cortical thickness. In contrast, trabecular size or thickness were less relevant. The pQCT-index regression model revealed total vBMD and bone volume fraction as protective factor, while bone separation as risk factor, among all features. Our findings were not consistent with previous report of Ct.vBMD, Tb.Th, and stiffness being best predictors^24^. This could be due to difference in patient demographics, bone types, or algorithms, and would require future validation in larger cohort of elderly postmenopausal Chinese women.

Identifying HR-pQCT patient subtypes was valuable for personalized management. In our analyses, cluster 1 patients had higher probability of having fragility fractures and osteoporosis by aBMD. Consistently, multiple aberrancies were found in HR-pQCT besides vBMD, including fewer trabecular number, higher trabecular separation, lower bone volume fraction, smaller cortical area, and lower cortical thickness. Of note, not all cluster 1 patients met diagnosis of osteoporosis by aBMD, and for those who did not, HR-pQCT phenotyping combined with DeepQCT scores would greatly facilitate early interventions, including weight-bearing exercise, vitamin D-calcium supplement, and intensive anti-osteoporosis treatment to prevent further progression to fracture events.

Our study had several limitations. First, prediction was restricted to history but not future events, due to current limitation of prospective data. However, fracture history is strongly relevant to future fracture, and our model can be easily adapted with upcoming data in ChiVOS^41^. Second, cohort size was limited compared to other international studies^26^. For this, we used augmentation for HR-pQCT images to increase size of training set to four times; and from performance, both DL and traditional logistic regression reached satisfactory AUC. Another issue is lack of external validation set, as a similar community cohort of same demographics was currently unavailable. We instead used an intact internal test set, independent of development set for cross validation to keep result integrity. Also, we here only included female of 50 years or older in current cohort, thus younger adults or male patients not covered. Nevertheless, the ChiVOS has been actively recruiting, and most aforementioned issues will most likely resolve in future studies. Importantly, the current methodology is fully applicable to new data.

In conclusion, we developed DeepQCT, a deep learning model which predicts fragility fracture history directly using HR-pQCT scans, and which outperformed BMD models in ChiVOS cohort. We identified a subset of microarchitectural parameters more closely related to vBMD, and revealed a patient subgroup with multiple bone density and microstructural abnormalities showing higher fragility fracture risk. Our findings established methodological framework of direct using HR-pQCT bone scans for risk prediction or diagnosis of bone mineral diseases, to facilitate early recognition and primary prevention in patients susceptible to osteoporotic fracture.

## Data Availability

Desensitized demographic data, HR-pQCT indices, HR-pQCT images, and codes of the current study would be available upon reasonable request to corresponding author.

## Contributors

LC and QJ developed concept and design of this study. FC performed data analysis, data interpretation, statistical analysis, and drafted the manuscript. YJ, WL, YC, RJ, QP, OW, ML, XX, WY, WX established the ChiVOS cohort used for data acquisition. LC, YW, and JL participated in data acquisition. JL, QJ, and XZ provided administrative, technical, or material support in data analysis. XZ and WX provided supervision. All authors contributed to critical revision of the manuscript for important intellectual content.

## Acknowledgements

This research was supported by the National Natural Science Foundation of China (LC, 82100946; WX, 82270938), CAMS Innovation Fund for Medical Sciences (WX, 2021-I2M-1-002), National Key R&D Program of China (WX, 2021YFC2501700), National High Level Hospital Clinical Research Funding (WX, 2022-PUMCH-D-006), the Non-profit Central Research Institute Fund of Chinese Academy of Medical Sciences (LC, 2023-PT320-10), and Young Elite Scientists Sponsorship Program by BAST (LC, No.BYESS2023171). Part of the study was supported by Merck Sharp & Dohme China, Hangzhou, China.

## Figure Legend

**Figure S1. ROC curves of all models predicting fragility fracture history** Receiver operating characteristic (ROC) curves of BMD model (A), pQCT-index models (B-D), and DeepQCT models (E-I), showing AUC, optimal cut-off value of predicted probability, and sensitivity and specificity at optimal cut-off at each cross-validation fold.

