## Supplementary figures and images for "DeepQCT: Predicting fragility fracture from high-resolution peripheral quantitative CT using deep learning"

### Supplementary Figures

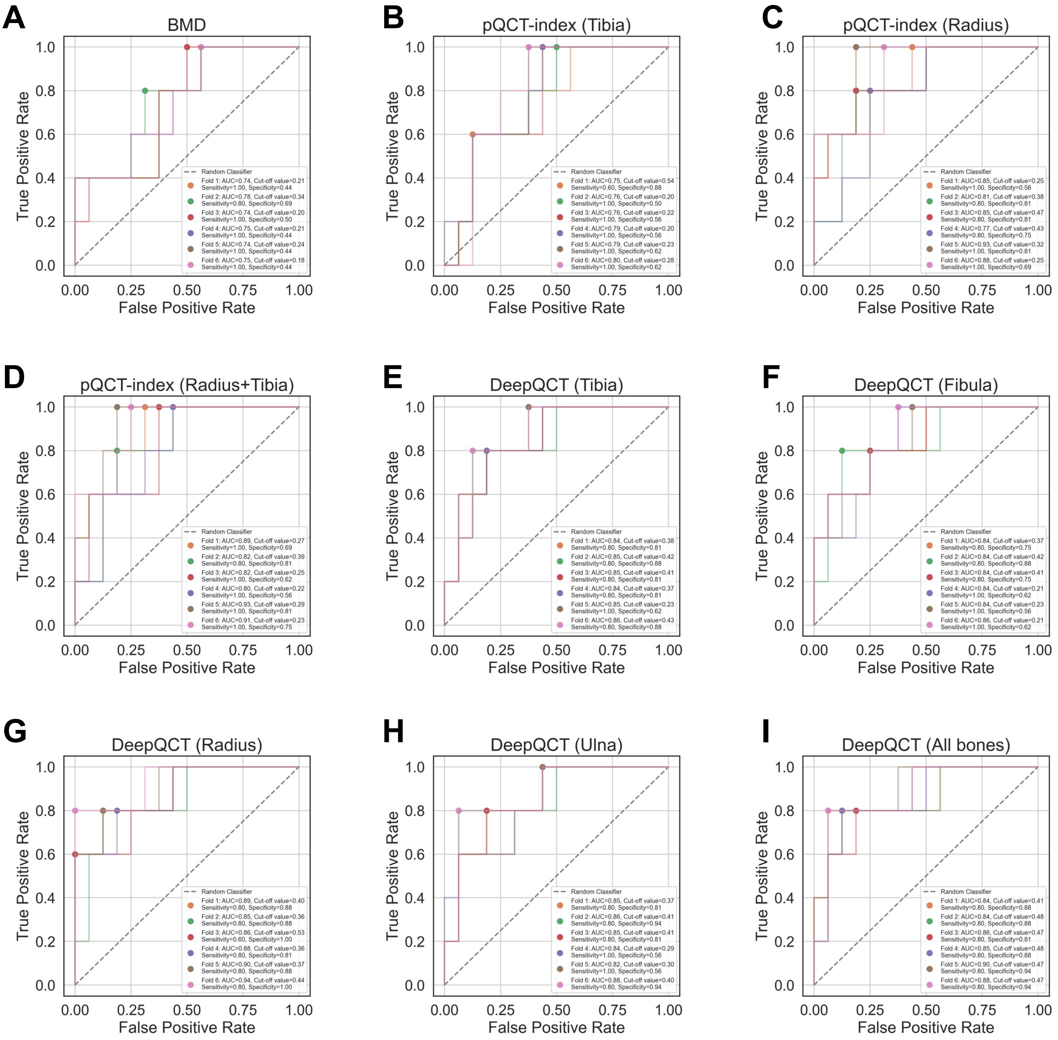
